# Efficacy of sovateltide (IRL-1620) in a multicenter randomized controlled clinical trial in patients with acute cerebral ischemic stroke

**DOI:** 10.1101/2020.08.17.20176784

**Authors:** Anil Gulati, Nilesh Agrawal, Deepti Vibha, U.K. Misra, Birinder Paul, Dinesh Jain, Jeyaraj Pandian, Rupam Borgohain

## Abstract

**Background:** Sovateltide (IRL-1620, PMZ-1620), an endothelin-B receptor agonist, administered intravenously following acute cerebral ischemic stroke increased cerebral blood flow, had anti-apoptotic activity and produced neurovascular remodeling. Its safety and tolerability were confirmed in healthy human volunteers (CTRI/2016/11/007509).

**Objective:** To determine safety, tolerability and efficacy of sovateltide as an adjuvant to standard of care (SOC) in acute cerebral ischemic stroke patients.

**Methods:** A prospective, multi-centric, randomized, double-blind, controlled study to compare efficacy of sovateltide in patients with acute cerebral ischemic stroke was conducted in 40 patients, of which 36 completed 90-day follow-up. Patients who had stroke within the last 24 hours with a radiologic confirmation of ischemic stroke were included in the study. Patients with intracranial hemorrhage and those receiving endovascular therapy were excluded. All patients in both groups received SOC for stroke. Patients randomized in the sovateltide group received three doses of sovateltide (each dose 0.3 µg/kg) administered as an IV bolus over 1 minute at an interval of 3 hours ± 1 hour on day 1, day 3 and day 6 (total dose of 0.9 µg/kg/day). Patients randomized in the placebo group received equal volume of saline. Efficacy was evaluated by neurological outcomes based on National Institute of Health Stroke Scale (NIHSS), modified Rankin Scale (mRS) and Barthel Index (BI) scales. Quality of Life was measured by EuroQoL (EQ-5D) and stroke specific quality-of-life (SS-QoL).

**Results:** Patients received saline (n = 18; 11 male and 7 female) or sovateltide (n = 18; 15 male and 3 female) within 24 hours of onset of stroke. Number of patients receiving investigational drug within 20 hours of onset of stroke were 14/18 in saline and 10/18 in sovateltide cohorts. The baseline characteristics and SOC in both cohorts was similar. Sovateltide treatment resulted in a significantly quicker recovery as measured by improvements in neurological outcomes in mRS and BI scales on day 6 compared to day 1 (p< 0.0001). Moreover, sovateltide increased the frequency of favorable outcomes in all scales at 3 months. An improvement of ≥2 points in mRS was observed in 60% and 40% patients in sovateltide and saline groups, respectively (p = 0.0519; odds ratio 5.25). BI improvement of ≥40 points was 64% and 36% in sovateltide and saline groups, respectively (p = 0.0112; odds ratio 12.44). An improvement of ≥6 points was seen in NIHSS in 56% of patients in sovateltide vs 43% in saline groups (p = 0.2714; odds ratio 2.275). Number of patients with complete recovery achieving NIHSS score of 0 and BI of 100 were significantly more (p< 0.05) in sovateltide group compared to saline group. Sovateltide treatment resulted in improved Quality of Life as measured by EuroQoL and SS-QoL (stroke specific quality-of-life) on day 90. Sovateltide was well tolerated and all patients received complete treatment with no incidence of drug related adverse event reported. Hemodynamic, biochemical or hematological parameters were not affected with sovateltide.

**Conclusion:** Sovateltide was safe, well tolerated, and resulted in quicker recovery and improved neurological outcome in acute cerebral ischemic stroke patients 90 days post-treatment.

**Trial Registration:** The study is registered at CTRI/2017/11/010654 and NCT04046484

**Key Points:** A phase II trial was conducted to evaluate safety, tolerability and efficacy of sovateltide, an ET_B_ receptor agonist, in patients with acute cerebral ischemic stroke.

Sovateltide was administered in three doses, each dose of 0.3 µg/kg, as an intravenous bolus over one minute at an interval of 3 hours ± 1 hour on day 1, day 3, and day 6 (total dose/day: 0.9 µg/kg) post randomization.

It was found to be safe and well tolerated. No adverse event was reported in patients.

Sovateltide significantly improved outcome parameters of NIHSS, mRS and BI at day 6 compared to day 1, however no significant improvement was observed in control arm.

Number of patients showing an improvement in mRS and BI were greater in sovateltide compared to control cohort at 90 days of treatment.

## 1. Introduction

Stroke is the fifth prominent cause of death in the USA. It is also an important reason for serious long-lasting disability of stroke patients. Ischemic stroke caused by arterial occlusion is responsible for most strokes [1]. The currently available treatment for acute cerebral ischemic stroke includes tissue plasminogen activator (tPA) and mechanical thrombectomy. However, the use of tPA is limited to a short time window of < 4.5 h from the onset of symptoms [2] and it also has a 4.9% risk of intracranial hemorrhage [3]. Mechanical thrombectomy was limited to 6 hours after onset of stroke symptoms has been extended up to 24 hours [4]. Limitations of time, risk of bleeding and involvement of larger blood vessels for thrombectomy restricts current treatment only to a handful (about 5%) of patients. Development of new effective drugs for acute cerebral ischemic stroke to alleviate neurological deficit and repair the cerebral damage is urgently required. Significant effort is being made to understand the complex pathophysiology of stroke and various mechanisms e.g. anticoagulation, neuroprotection and neuroregeneration are being explored [5–8]. Despite many encouraging preclinical results, numerous drugs as potential neurovascular protectants failed to demonstrate their beneficial effects in clinical trials [6, 9]. One of the reasons of these failures could be the complex pathophysiology of ischemic stroke, which involves hypoxia, vascular damage, inflammation, apoptosis and other events leading to neural cell damage and functional impairment of the brain. Therefore, a new approach is needed which has the potential to address the preservation of salvageable brain tissue, minimizing complications, and secondary prevention.

Levels of ET-1 have been reported to be elevated in the blood and brain tissues following cerebral ischemia [10, 11]. Since ET-1 has been described as a potent vasoconstrictor via acting on endothelin A (ET_A_) receptors, it was hypothesized that ET_A_ receptor antagonists would reduce the damage associated with acute cerebral ischemic stroke. Studies focused on antagonizing ET_A_ receptors using BQ123, SB234551, A-127722 and S-1039, demonstrated a reduction in infarct area, edema, and neurological deficit following experimental cerebral ischemia [12–16] but have not advanced to clinical stage. Combined ET_A/B_ receptor antagonists have shown mixed results, TAK-044 decreased oxidative stress and reduced ischemia, while bosentan and SB209670 had no effect [17, 18]. On the contrary, antagonizing ET_B_ receptors worsens ischemic injury, leading to poor outcomes [19, 20], suggesting critical role of ET_B_ receptors in salvaging brain damage due to cerebral ischemia.

ET_B_ receptors are present in large numbers in the central nervous system (CNS) and play a key role in its development. We have demonstrated that ET_B_ receptors in the brain are over expressed at the time of birth and their expression decreases with maturity of the brain [21, 22], suggesting that they are important in the brain development and may be useful in repair and regeneration of adult brains after stroke [5, 23, 24]. ET_B_ receptors play an important role in the CNS development, neural cell survival and proliferation [5, 25–28]. Stimulation of ET_B_ receptors in middle cerebral artery occluded (MCAO) rats with intravenous administration of sovateltide, a highly selective ET_B_ receptor agonist, significantly improved neurological and motor functions, decreased infarct volume, oxidative stress and apoptosis damage, and increased cell proliferation and angiogenesis [24, 29, 30]. Sovateltide induced improvements were blocked with ET_B_ receptor antagonist, BQ788, confirming ET_B_ receptor involvement in sovateltide effects [29, 30].

Sovateltide demonstrated potential to be developed as a novel and effective drug to treat acute cerebral ischemic stroke. Safety and tolerability of sovateltide through clinical (CTRI/2016/11/007509) phase I study in healthy human volunteers has been demonstrated [5]. The present study is presenting clinical evidence of leveraging developmental mechanism by stimulating ET_B_ receptors by sovateltide to ameliorate and restore nervous system functions following cerebral ischemic stroke in adult patients.

## 2. Methods

This was a prospective, multi-center, randomized, placebo-controlled, double-blinded phase 2 clinical study to evaluate the safety and efficacy of sovateltide as an adjuvant to standard of care (SOC) in patients with acute ischemic stroke. Primary objective of the study was to evaluate the safety and tolerability of sovateltide. Key secondary objectives included efficacy measurements of neurological improvements by NIHSS, mRS and BI scales and quality of life assessments by Euro Qol and stroke-specific quality of life (SSQoL).

### 2.1. Patients, Eligibility Criteria and Trial Design

Key inclusion criteria were the following: age from 18 years through 70 years, signed informed consent by patient or through a legally authorized representative (LAR) if the patient is not in a condition to give consent, onset of stroke symptoms within last 24 hours with mRS score of 3–4 and NIHSS score of 5–14, stroke is ischemic in origin, supratentorial and radiologic confirmation either with CT scan or diagnostic MRI and patients receiving thrombolytic therapy were included in the study. Female patients were either of not child-bearing potential or on double-contraception. Key exclusion criteria included the following: patients receiving endovascular therapy, those presenting with intracranial hemorrhage, recurrent stroke, participating in other therapeutic clinical trial, evidence of major life-threatening or serious medical condition and pregnant, breast feeding women or positive pregnancy test.

The study was done in accordance with ICH-GCP Guidelines, the principles of the Declaration of Helsinki and local regulatory requirements. The study protocol (PMZ-01 Version 2.0/April 18, 2016) was approved by the Drugs Controller General of India (DCGI), Directorate General of Health Services, Ministry of Health and Family Welfare, Government of India and Institutional Ethics Committee of each of 6 sites reviewed and approved the study protocol before the site was initiated. The study was registered at the Clinical Trials Registry, India (CTRI/2017/11/010654) and NCT04046484. Ethics committee of each site was continuously informed of any protocol deviation, amendments, subject exclusion or withdrawal and serious adverse events. Informed consent was obtained from each patient, those patients who were not fit to give consent themselves at the time of initiation of treatment, informed consent was taken from their legally authorized representative (LAR). The patient/LAR was informed by the investigator in writing with audio-visual recording about all aspects of the study relevant to taking a decision on whether to participate in the study or not. The informed consent form included all the elements required as per the ICH-GCP recommendations and schedule Y.

### 2.2. Treatment Regimen

Patients were randomized 1:1 either to the sovateltide group or the placebo group. Block randomization was used for patient randomization into the 2 treatment groups. The randomization list was prepared by a statistician using a validated computer program, statistical analysis system SPSS. An Interactive Web Response System (IWRS) method containing randomization codes was used to randomize the eligible patient to the treatment groups.

All patients in both groups received SOC for stroke according to their local institutional guidelines. Patients in the sovateltide group received three doses of sovateltide (each dose 0.3 µg/kg) administered as an IV bolus over 1 minute at an interval of 3 hours ± 1 hour on day 1, day 3 and day 6 (total dose of 0.9 µg/kg/day). Patients in the placebo group received equal volume of saline as an IV bolus over 1 minute at an interval of 3 hours ± 1 hour on day 1, day 3 and day 6. Duration of the study for each patient was 3 months (90 days) which included 5 study visits: visit 1/Day 1 (screening/baseline measurements/treatment, visit 2/Day 12, visit 3 (Day 30 ± 5 days, visit 4 (Day 60 ± 5 days and visit 5/End of study (Day 90 ± 5 days). All patients were monitored closely throughout hospitalization for qualifying stroke and followed-up for 90 days from randomization.

### 2.3. Data Safety Monitoring Board

An independent Data Safety Monitoring Board consisting of a neurologist, a biostatistician, and a clinical pharmacologist was established to monitor the safety and efficacy of the trial. The Data Safety Monitoring Board reviewed safety data of each subject from the study and reviewed all serious adverse events, regardless of attribution, contemporaneously with submissions to the sponsor and investigator.

### 2.4. Safety and Tolerability Assessments

All patients who received treatment were included in safety analysis. Safety was assessed during treatment and post-treatment follow-up period based on adverse events, physical examination, vital signs and clinical laboratory parameters as per protocol. A variety of biochemical tests, serum chemistry tests, hematological variables and organ function tests such as kidney and liver function were performed. Adverse events that occurred or worsened during treatment or post-treatment were recorded. All AEs were coded by preferred term and system organ class using the latest version of MedDRA. All patients were followed up for safety assessment till the end of study on Day 90.

### 2.5. Assessment of Neurological Outcome

Efficacy of sovateltide was evaluated by three different outcome measures. Proportion of patients with change in NIHSS, mRS and BI scores was evaluated on day 6, 1 month and 3 months post-randomization. Improvements of ≥ 6 points in NIHSS over the baseline score was considered as a favorable outcome. Similarly, improvement of ≥ 2 points in mRS and improvement of ≥ 40 points in BI scale, from the baseline score was considered as a favorable outcome. Proportion of patients with overall clinical improvements was assessed by global statistical test of NIHSS, mRS and BI scores at 3 months post-randomization. Quality of life assessments were performed by validated method of EuroQol and stroke-specific quality of life (SSQOL) at 3 months post-randomization. In the EuroQol, a score of 100 was defined as the best health for the patient. Similarly, in the SSQOL a score of more than 200 was defined as the best health for the patient.

### 2.6. Blinding/Unblinding

In this double-blind study, the patient and all relevant personnel involved with the conduct and interpretation of the study (including investigator, investigational site personnel, and the sponsor or designee’s staff) were blinded to the identity of the study drug (sovateltide/normal saline) assigned and the randomization codes. The biostatistician/unblinded pharmacist was independent of study team. Dispensing activity was monitored by an unblinded monitor independent of monitoring team. Final randomization list was kept strictly confidential and accessible only to authorized persons per sponsor until the completion of the study.

Emergency unblinding through IWRS was available. As per the study protocol, the investigator or his/her designee was permitted to unblind the code when medically needed, without identifying other patient’s treatment. For those patents, where unblinding was done, the date, time and the reason for emergency unblinding was recorded in patient’s medical record. Any AE or SAE that required unblinding the treatment was recorded and reported as specified in the protocol. Treatment unblinding was not done for any of the patients enrolled in this study.

### 2.7. Sample Size & Statistical Methods

Data obtained from earlier clinical studies of stroke in the literature was considered for sample size determination for this phase 2 study. Assuming 80% power and 5% significance level, a sample size of 32 patients (16 patients in each group) was required. Considering a 10% loss to follow-up, the final sample size was 36 patients was required. To increase the power of the study, we decided to increase the sample size to 40 patients (20 patients in each group).

The results of the trial are presented as mean ± SEM. The significance of differences was estimated by Tukey’s multiple comparisons test, Chi-square test and Unpaired t test with Welch’s correction. A P value of less than 0.05 was considered to be significant. The continuous variables between the groups were compared by using Unpaired t-test. Demographic variables and patient characteristics were summarized descriptively by treatment assignments. Demographic variables include age, weight, height, and body mass index. Variables that are measured on a continuous scale, such as the age of the patient at the time of enrolment, the number of non-missing observations (n), mean and SEM were tabulated by treatment assignments. The Unpaired t-test was used to compare the discrete variables between the groups at baseline and at follow-ups. All available data was used in the analyses. Each group was summarized individually. Data not available was assessed as “missing values” and the observed population only were evaluated. The statistical analysis was processed with GraphPad Prism 8.1.1 (GraphPad, San Diego, CA).

## 3. Results

### 3.1. Demographics and Patient characteristics

A total of 516 patients were assessed for the study in 6 clinical sites across India, out of which 40 patients met the eligibility criteria. To be eligible for this study, patients were to be diagnosed with stroke for the first time. Majority of patients screened for this study had a recurrent stroke, hence they were considered screen failures and they were not randomized for the study treatment. All 40 eligible patients signed informed consent and were randomized to either placebo (n = 20) or sovateltide groups (n = 20). All patients in both groups received SOC according to local institutional guidelines (Table 1). Out of 40 patients, 36 completed the study with a 90-day follow-up. In the control group, 18 patients completed the study. (1 patient withdrew consent and 1 patient was withdrawn by the investigator). Similarly, in the sovateltide group 18 patients completed the study (1 patient withdrew consent and 1 patient was withdrawn by the investigator (Figure 1). Patient demographics and baseline characteristics including baseline scores of NIHSS, mRS and BI were comparable in both groups (Table 2). Sovateltide group had more male patients than females (15M/3F) compared to placebo group (11M/7F). Age, body weight, BMI were similar in both groups.

**Figure 1:**
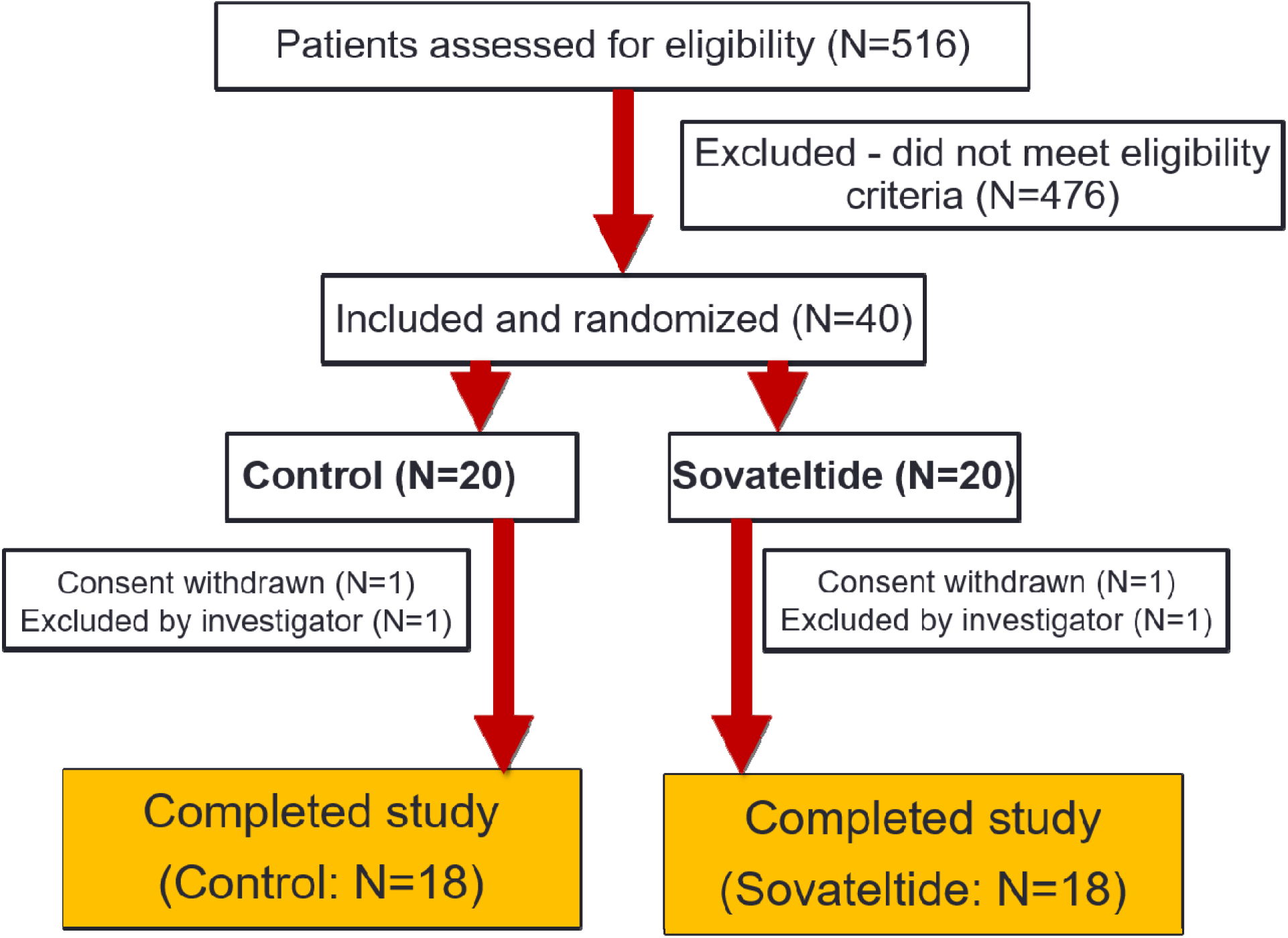
Screening and enrollment of patients

**Table 1:** Sites participating in the study

| **S. No.** | **Study Site** | **Total Enrolled** |
| --- | --- | --- |
| **1** | NewEra Hospital, Near Jalaram Mandir, Queta Colony, Telephone Exchange Chowk, Central Avenue Road, Nagpur 440008 Maharashtra | 12 |
| **2** | Christian Medical College and Hospital, Department of Neurology, Brown Road, Ludhiana -141008 Punjab | 02 |
| **3** | All India Institute of Medical Sciences, Department of Neurology, Neurosciences Centre, Ansari Nagar, New Delhi -110029 | 08 |
| **4** | Sanjay Gandhi Post Graduate Institute of Medical Sciences, Department of Neurology, Ground Floor, Raebareli Road, Lucknow- 226014 Uttar Pradesh | 05 |
| **5** | Dayanand Medical College & Hospital, Tagore Nagar, Civil Lines, Ludhiana, 141001 Punjab | 10 |
| **6** | Nizam's Institute of Medical Sciences. Punjagutta, Hyderabad 500082, Telangana | 02 |

**Table 2:** Demographics of patients in standard of care plus saline and standard of care plus sovateltide cohorts.

| **Patient Demographics (Mean ± SEM)** |  |  |  |  |  |
| --- | --- | --- | --- | --- | --- |
| **Group** | **Gender** | **Age (Years)** | **Body Weight (Kg)** | **Height (Cm)** | **BMI (Kg/m<sup>2</sup>)** |
| Normal Saline (N=18) | 11M/7F | 58.34±1.73 | 64.97±2.40 | 163.72±2.08 | 24.47±0.98 |
| Sovateltide (N=18) | 15M/3F | 49.83±2.79 | 67.82±2.47 | 168.78±1.50 | 24.00±0.68 |

All patients enrolled in the study (irrespective of the study group) received SOC treatment for ischemic stroke. Sovateltide was administered as an add-on to the SOC treatment. SOC treatment in both groups was similar and the details of the treatment are given in Tables 3A and 3B. All patients received the study drug within 24 hours of onset of stroke. However, more patients received treatment within 20 hours in the control group compared to sovateltide group where more patients received treatment after 20 hours. In the control group, 77.78% of patients received the treatment within 20 hours (14 out of 18 enrolled patients) and 22.22 % received treatment after 20 hours (4 out of 18 patients) while in the sovateltide group, 55.56% patients received treatment (SOC plus sovateltide) within 20 hours (10 out of 18 enrolled patients) and 44.45% patients received treatment after 20 hours (8 out of 18 enrolled patients) of stroke onset (Figure 2).

**Figure 2:**
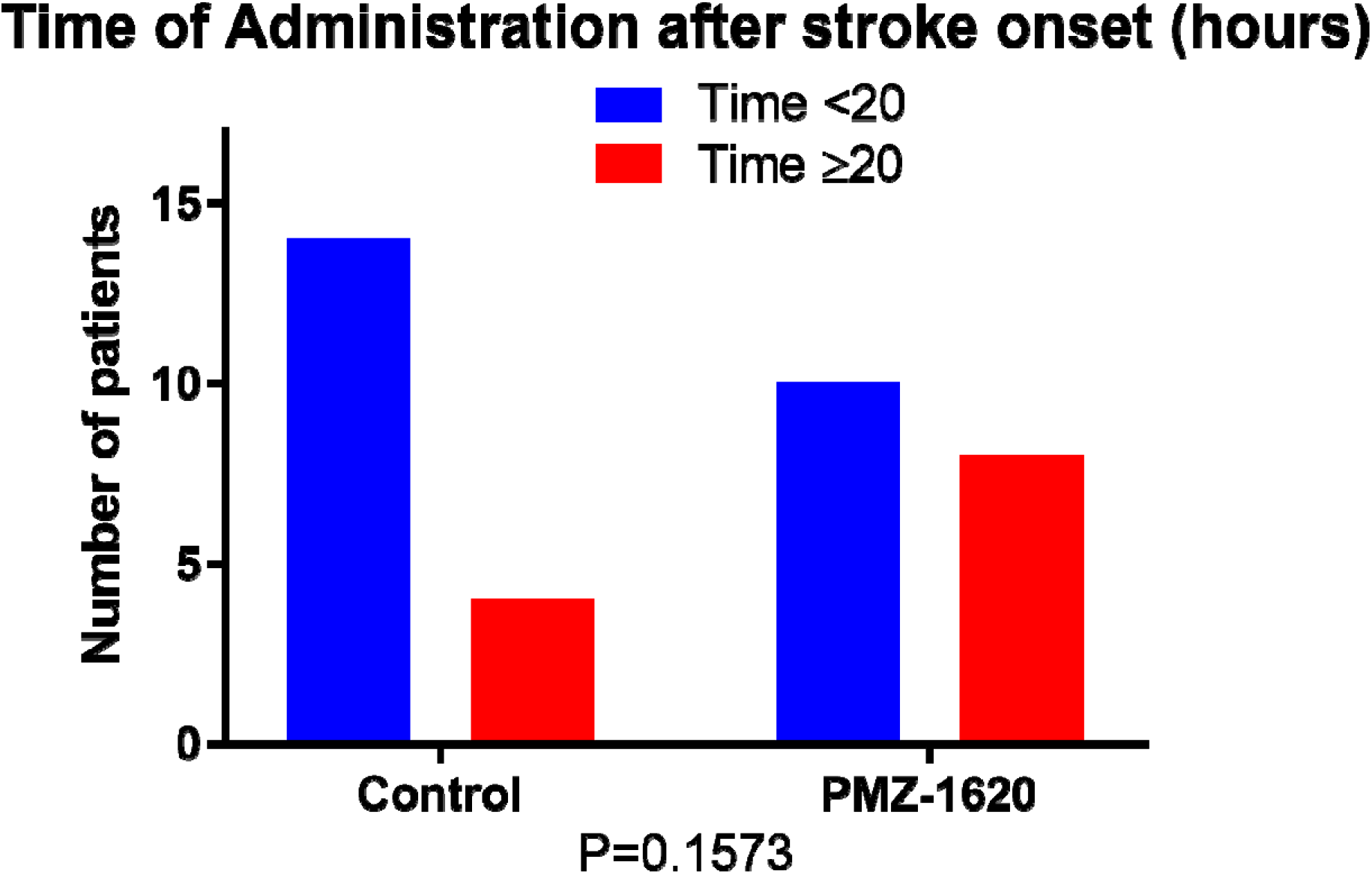
Time of enrollment and initiating the treatment after onset of stroke.

**Table 3A:** Standard of care details of the treatment in control group of patients

| Standard Treatment + Saline (N=18) |  |  |
| --- | --- | --- |
| S. No | Patient Number | Treatment |
| 1 | 01-001 | Inj. Enoxaparin, Tab. Aspirin, Tab Rosuvastatin, Tab. Cilnidipine and Chlorthalidone |
| 2 | 01-004 | Inj. Enoxaparin, Tab. Aspirin, Tab. Clopidogrel and Tab. Rosuvastatin |
| 3 | 01-005 | Inj. Cognistar, Tab Levetiracetam, Inj. Mannitol, Tab. Rosuvastatin, Tab. Amlodipine, Tab. Aspirin and Tab. Furosemide Rosuvastatin |
| 4 | 01-008 | Inj. Enoxaparin, Tab. Aspirin & Clopidogrel and Tab. Rosuvastatin |
| 5 | 01-009 | Tab. Aspirin & Clopidogrel, Tab. Atorvastatin and Inj. Cognistar |
| 6 | 01-012 | Inj. Enoxaparin, Inj. Cognistar, Tab. Aspirin & Clopidogrel, Tab. Rosuvastatin and Tab. Amlodipine |
| 7 | 03-001 | Cerebral decongestants, Tab. Aspirin, Tab. Amlodipine and Tab. Atorvastatin |
| 8 | 03-003 | Tab. Aspirin and Tab. Atorvastatin |
| 9 | 03-006 | Tab. Amlodipine, Tab. Aspirin, Tab. Clopidogrel and Tab. Atorvastatin |
| 10 | 03-008 | Tab. Amlodipine, Tenecteplase, Tab. Aspirin and Tab. Atorvastatin |
| 11 | 04-002 | Cap. Aspirin and Atorvastatin |
| 12 | 04-003 | Angiogenic medication, Cap. Aspirin & Atorvastatin and Tab. Clopidogrel |
| 13 | 04-005 | Tab. atorvastatin, Tab. Aspirin, Tab. Telmisartan and Tab. Clopidogrel |
| 14 | 06-002 | Tab. Aspirin & Clopidogrel, Inj. Mannitol, Tab. Atorvastatin and Inj. Dalteparin |
| 15 | 06-006 | Tab. Aspirin, Tab. Atorvastatin and Tab. Telmisartan and Amlodipine |
| 16 | 06-007 | Tab. Aspirin, Tab Atorvastatin, Tab. Amlodipine, Tab. Telmisartan and Cap. Fluoxetine |
| 17 | 06-009 | Tab. Telmisartan & Amlodipine, Tab. Aspirin, Atorvastatin & Clopidogrel and Tab. Memantine |
| 18 | 07-002 | Tab. Aspirin and Tab. Atorvastatin |

**Table 3B:** Standard of care details of the treatment in sovateltide group of patients

| Standard Treatment + Sovateltide (N=18) |  |  |
| --- | --- | --- |
| S. No | Patient Number | Treatment |
| 1 | 01-002 | Inj. Enoxaparin, Tab. Rosuvastatin, Tab. Amlodipine, Tab. Aspirin and Clopidogrel |
| 2 | 01-003 | Tab. Amlodipine, Inj. Enoxaparin, Inj. Cognistar, Tab. Aspirin, Clopidogrel, Tab. Telmisartan and Tab. Rosuvastatin |
| 3 | 01-006 | Inj. Enoxaparin, Tab. Aspirin & Clopidogrel and Tab. Amlodipine |
| 4 | 01-007 | Inj. Enoxaparin, Tab. Aspirin & Clopidogrel, Tab Rosuvastatin, Inj. Cognistar, Tab. Labetalol, Tab. Amlodipine and Tab. Nifedipine |
| 5 | 01-010 | Inj. Enoxaparin, Tab. Aspirin & Clopidogrel and Inj. Mannitol |
| 6 | 01-011 | Tab. Atorvastatin, Inj. Enoxaparin, Inj. Cognistar and Tab. Nicoumalone |
| 7 | 02-001 | Inj. Alteplase, Tab. Aspirin, Tab. Atorvastatin, Tab. Escitalopram, Tab. Telmisartan |
| 8 | 02-002 | Inj. Heparin, Tab. Aspirin, Tab. Atorvastatin and Inj. Enoxaparin |
| 9 | 03-004 | Tab. Amlodipine, Alteplase, Tab. Aspirin and Tab. Atorvastatin |
| 10 | 03-005 | Tab. Metoprolol, Tab. Aspirin, Tab. Clopidogrel, Tab. Ramipril and Tab. Atorvastatin |
| 11 | 03-007 | Tab. Atorvastatin, Tab. Aspirin, Tab. Clopidogrel, Tab. Fluoxetine and Tab. Amlodipine |
| 12 | 04-001 | Cap. Aspirin and Atorvastatin |
| 13 | 04-004 | Tab. Nicoumalone, Tab. Furosemide & Spironolactone and Inj. Enoxaparin |
| 14 | 06-001 | Tab. Aspirin, Tab. Clopidogrel, Tab. Atorvastatin and Tab. Trimetazidine |
| 15 | 06-004 | Tab. Aspirin and Tab. Atorvastatin |
| 16 | 06-005 | Tab. Aspirin, Inj. Mannitol, Inj. Enoxaparin, Inj. Furosemide and Tab. Atorvastatin |
| 17 | 06-008 | Tab. Olmesartan & Amlodipine, Tab. Atorvastatin, Syp. Phenytoin, Tab. Aspirin & Clopidogrel |
| 18 | 07-001 | Tab. Aspirin and Tab. Atorvastatin |

### 3.2. Significantly quicker recovery in patients treated with sovateltide

Neurological outcome measures in individual patients from day 1 through day 90, using three different scales are presented in Figure 3. In panel A, NIHSS score in individual patients on day 1 (baseline) and day 2, day 3, day 6, day 30, day 60, day 90 post randomization, is presented for both control and sovateltide treated groups. Data indicates trend of early recovery of patients from sovateltide group compared to control as seen from significant improvement in NIHSS score (sovateltide: day 3 NIHSS; p< 0.0001 vs Control group: day 3 NIHSS; p = 0.5771). In panel B, mRS score in individual patients on day 1 (baseline) and day 2, day 3, day 6, day 30, day 60, day 90 post randomization, is presented for both control and sovateltide treated groups. Data indicates trend of early recovery of patients from sovateltide group compared to control as seen from significant improvement in mRS score (sovateltide: day 6 mRS; p< 0.0001 vs Control group: day 6 mRS; p = 0.0859). In panel C, BI score in individual patients on day 1 (baseline) and day 2, day 3, day 6, day 30, day 60, day 90 post randomization, is presented for both control and sovateltide treated groups. Data indicates trend of early recovery of patients from sovateltide group compared to control as seen from significant improvement in BI score (sovateltide: day 6 BI; p< 0.0001 vs Control group: day 6 BI; p = 0.3948). Neurological outcome measures on day 1 versus day 6 in NIHSS scores are presented in Figure 4. An improvement of ≥6 points in NIHSS from baseline was seen in 87.50% of patients treated with sovateltide compared to 12.50% of patients in the control group (P = 0.0201). This data further supports the conclusion that patients treated with sovateltide recovered much quicker compared to patients treated with placebo.

**Figure 3:**
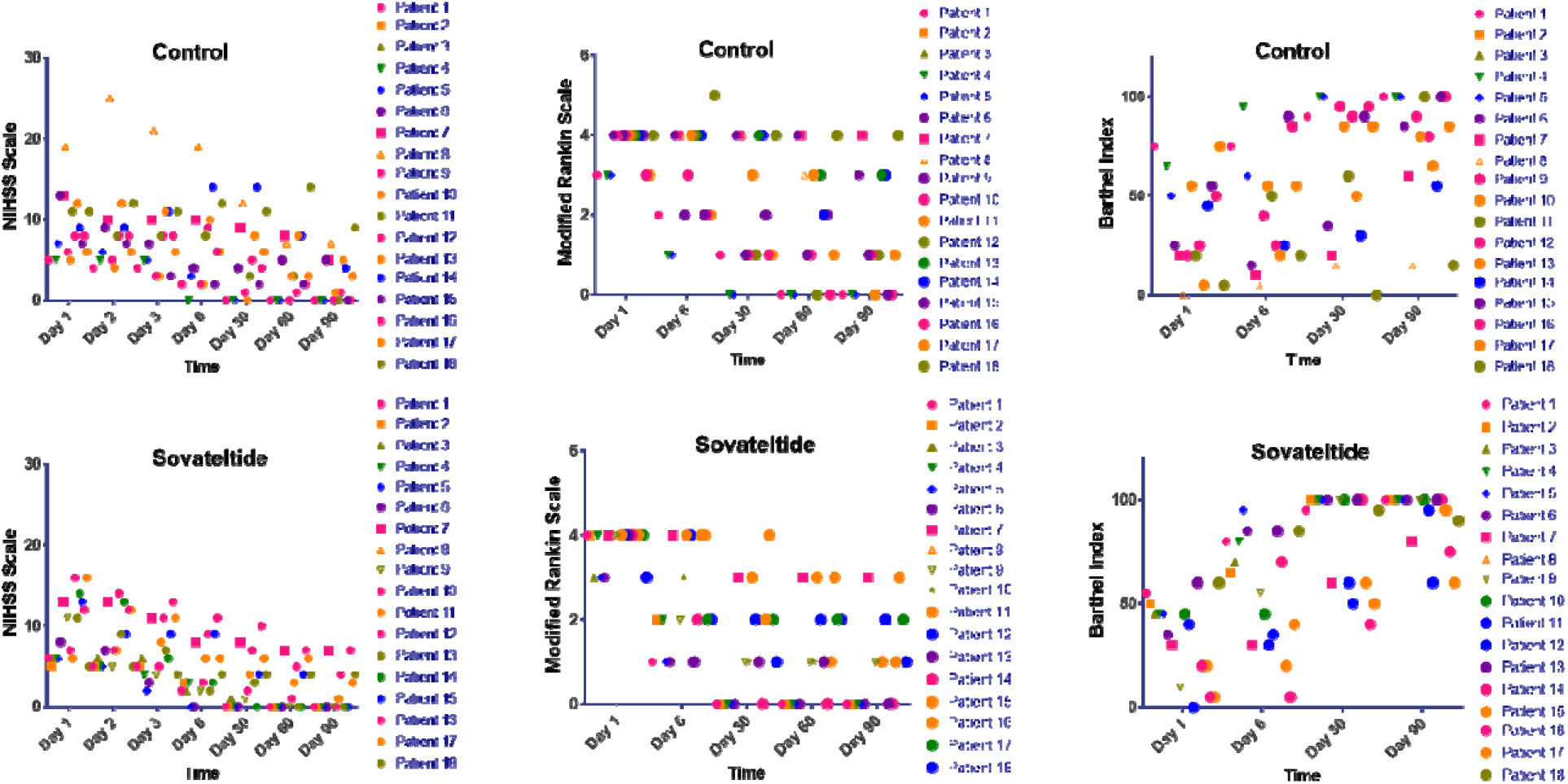
Neurological outcome measures of NIHSS, mRS and BI in individual patients from day 1 through day 90.

**Figure 4:**
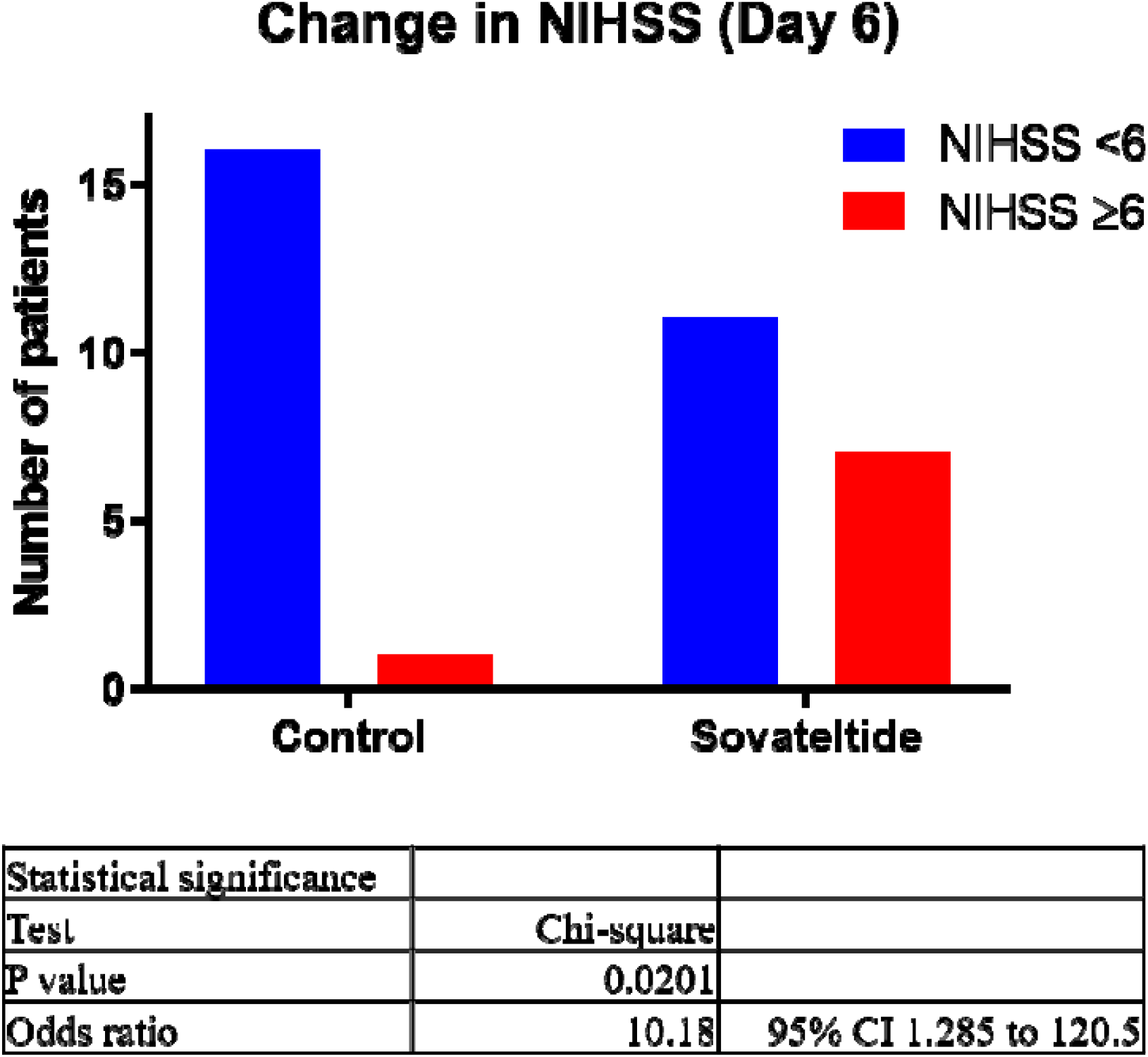
Neurological outcome measures on day 1 versus day 6 of NIHSS scores.

### 3.3. Sovateltide treatment increased the frequency of favorable outcomes at 3 months

Neurological outcomes at 3 months were measured using three different scales. Improvements of ≥ 6 points in NIHSS over the baseline score was considered as a favourable outcome. Similarly, improvement of ≥ 2 points in mRS and improvement of ≥ 40 points in BI scale, from the baseline score was considered as a favourable outcome. Sovateltide increased the frequency of favorable outcomes in all three scales at 3 months. An improvement of ≥2 points in mRS was observed in 60% and 40% patients in sovateltide and saline groups, respectively (p=0.0519; odds ratio 5.25). BI improvement of ≥40 points was 64% and 36% in sovateltide and saline groups, respectively (p = 0.0112; odds ratio 12.44). An improvement of ≥ 6 points was seen in NIHSS in 56% of patients in sovateltide vs 43% in saline groups (p = 0.2714; odds ratio 2.275) (Figure 5). Although statistical significance was not seen in NIHSS, the trend clearly shows an improvement in favorable outcome.

**Figure 5:**
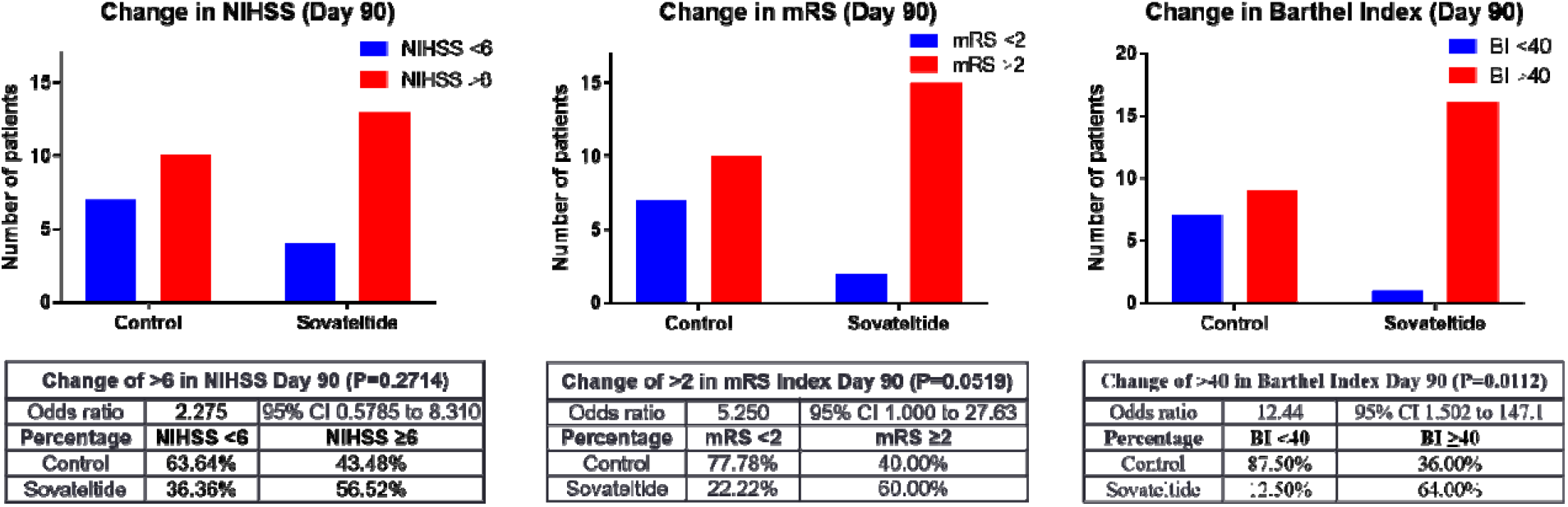
Number of patients with change in NIHSS, mRS and BI at 90 day of treatment.

### 3.4. Sovateltide treatment increased frequency of complete recovery

Comprehensive analysis of complete recovery was assessed using three different outcome scales in patients from control and sovateltide groups (Figure 6). Complete recovery was defined as a patient having NIHSS score of 0, mRS score of 0 and Barthel Index score of 100. Analysis of data shows that, sovateltide group patients had higher number of patients with NIHSS score of 0 (p = 0.04791). Sovateltide group also had higher number of patients with a mRS score of 0 (p = 0.1193). Similarly, sovateltide group patients had higher number of patients with a BI score of 100 counts indicating that sovateltide increased the frequency of complete recovery compared to control group patients (p = 0.02795). In another analysis, complete recovery was defined as patients with NIHSS score of 0–1, mRS score of 0–1 and BI score of 95–100. This is similar to the favourable outcome defined in the landmark rtPA 1195 stroke study. In this analysis also there was a clear trend towards complete recovery in the sovateltide group compared to control group, although it did not reach the level of statistical significance, however odds ratio was about 2.0. In the sovateltide group, 66% of patients had NIHSS score of 0–1 compared to 50% in the control group (odds ratio 2.00, 95% CI: 0.506–6.965, p value = 0.3105). In the sovateltide group, 72% of patients had mRS score of 0–1 compared to 55% in the control group (odds ratio 2.08, 95% CI: 0.484–8.476, p value = 0.2979). Similarly, in the sovateltide group, 66% of patients had BI score of 95–100 compared to 33% in the control group (odds ratio 4.00, 95% CI: 1.00–16.05, p value = 0.0455). These results clearly show a trend towards complete recovery in the sovateltide group compared to the control group.

**Figure 6:**
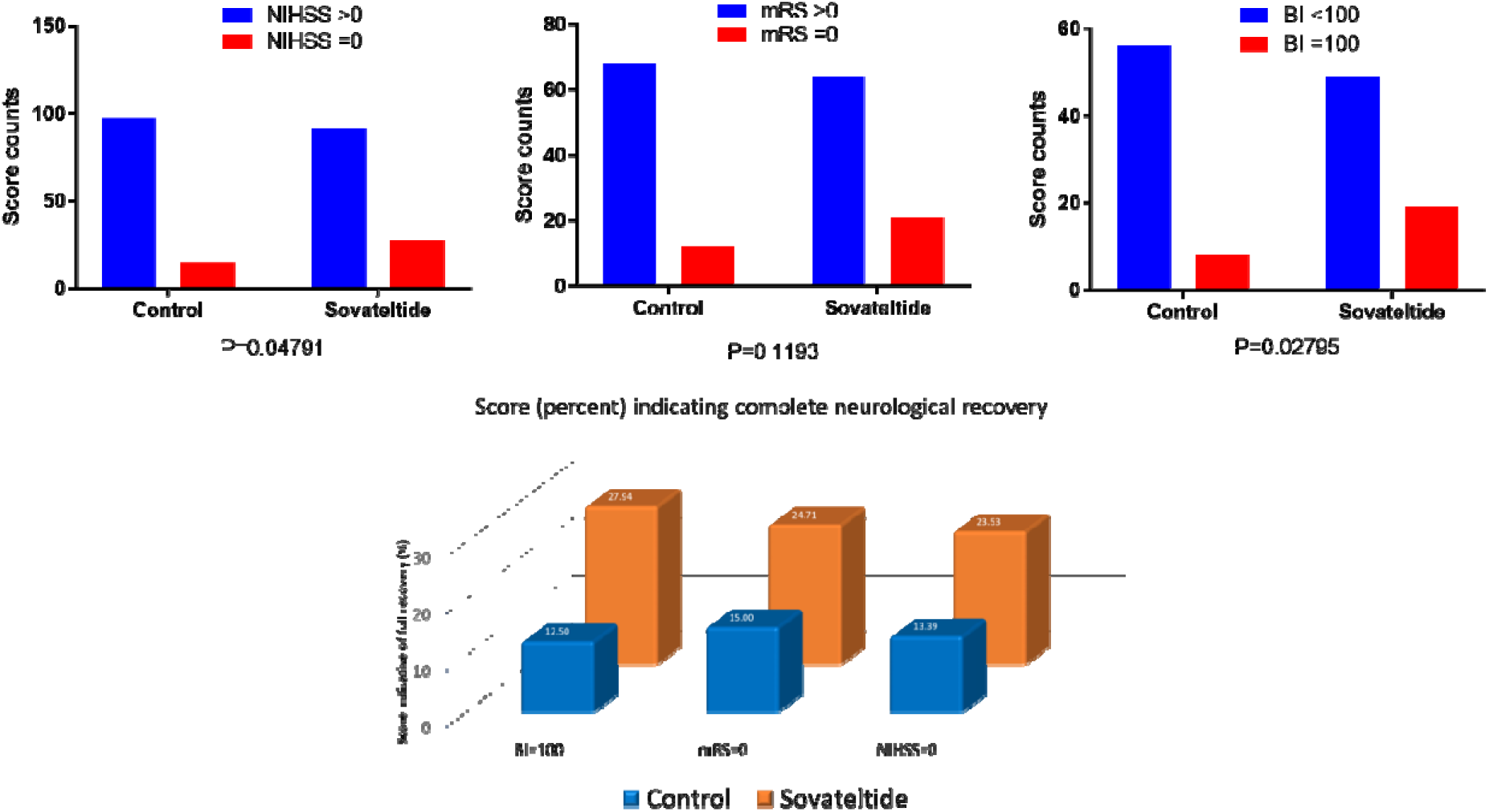
Number of patients with complete recovery as indicated by NIHSS, mRS and BI at 90 day of treatment.

### 3.5. Sovateltide treatment resulted in improved Quality of Life

Quality of life was assessed by two different scales, EuroQol and SSQoL (stroke specific quality of life). In the EuroQol, a score of 100 was defined as the best health for the patient. Similarly, in the SSQOL a score of more than 200 was defined as the best health for the patient. Both these measurements were done on day 60 and day 90. Statistical analysis of EuroQol data on day 60 showed odds ratio 3.250; outcome in sovateltide group is 225% likely to be better than control group. The 95% confidence interval for this odds ratio is between 0.7181 and 11.52. EuroQol data on day 90 showed odds ratio 1.956; outcome in sovateltide group is 95% likely to be better than control group. The 95% confidence interval for this odds ratio is between 0.4213 to 8.509 (Figure 7). Statistical analysis of SSQoL data on day 60 showed odds ratio 1.429; outcome in sovateltide group is 42% likely to be better than control group. The 95% confidence interval for this odds ratio is between 0.3565 and 4.907. SSQoL data on day 90 showed odds ratio 3.429; outcome in sovateltide group is 242% likely to be better than control group. The 95% confidence interval for this odds ratio is between 0.7909 to 15.47 (Figure 7).

**Figure 7:**
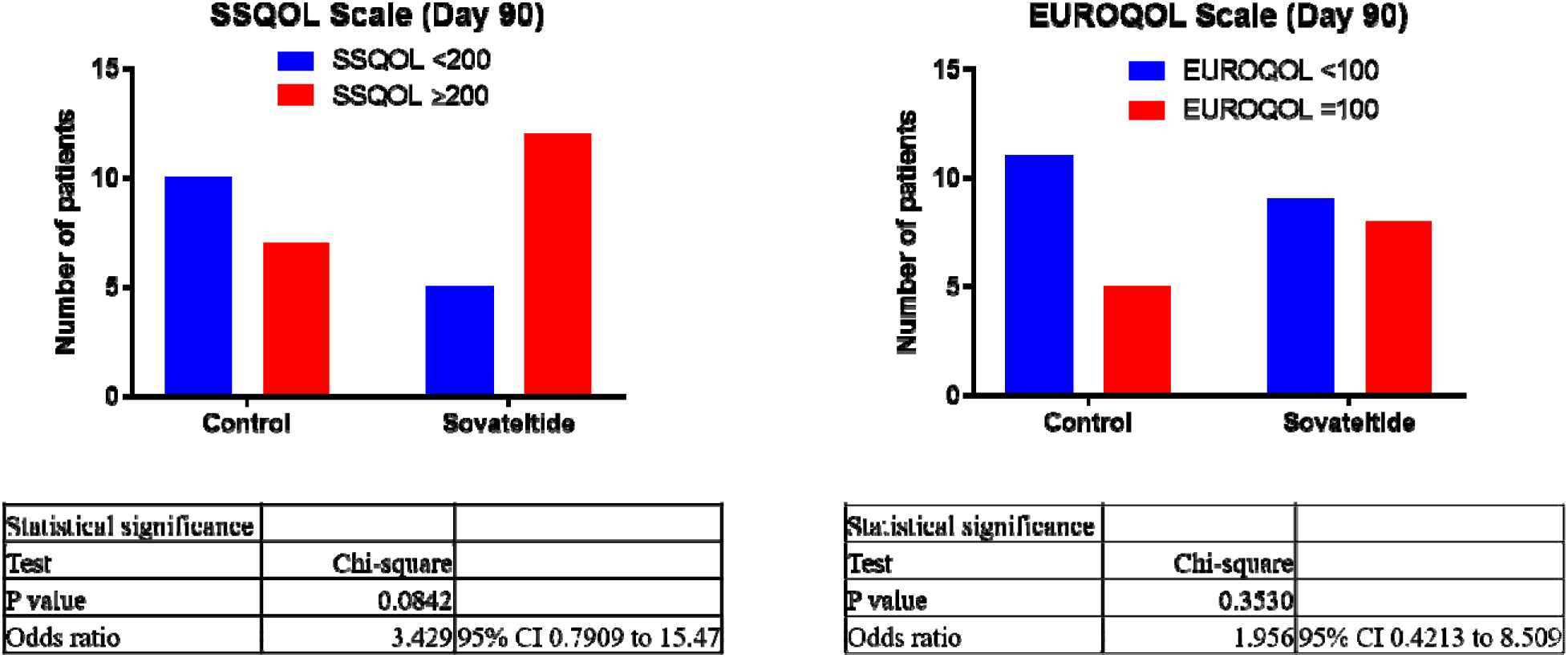
Number of patients with improved quality of life at 90 days of treatment.

### 3.6. Sovateltide is safe and well tolerated

Every patient enrolled received all 9 doses of sovateltide and none of the patient had any resistance to receiving the drug. All patients who received treatment were included in safety analysis. Sovateltide treatment did not have any effect on hemodynamic, biochemical or hematological parameters. No incidence of drug related adverse event was reported. There was no mortality and no incidence of recurrent ischemic stroke occurred in any of the enrolled patient. Sovateltide was found to be safe and was well tolerated in patients of ischemic stroke.

## 4. Discussion

Rapid restoration of blood flow by removing or dissolving thrombus from blood vessels is considered as the primary intervention of choice. Current available treatments, tPA and mechanical thrombectomy are categorized as primary intervention [31]. These primary interventions are beneficial only if provided quickly after the onset of stroke. For instance, administration of tPA after 4.5 hours of onset of stroke has shown no or poor effect in patients [32] and it can produce hemorrhagic transformation causing highly significant morbidity and mortality [32]. Attempts to improve recanalization of larger arteries following tPA by combining it with antiplatelet drugs have been unsuccessful with no change in 90 day clinical outcome [33, 34]. Granulocyte colony stimulating factor enhanced recanalization of the artery in patients with acute cerebral ischemic stroke without increasing hemorrhagic transformation with good neurological outcome 90 days post-treatment [35] and needs to be explored further. Sonothrombolysis as an adjuvant to intravenous thrombolysis has not shown any clinical benefit [36]. Hence, development of more potent and effective drugs is required to treat ischemic stroke.

Stroke pathophysiology involves hypoxia, vascular damage, inflammation, apoptosis and other events that severely affect neuronal cells [37]. Neuronal and glial cells are major components of neural tissues in the CNS and both are generated from a common Nestin expressing (Nestin+) neural stem/progenitor cells (NPCs) [38, 39]. Investigating a target protein or receptor in NPCs, which can generate new neuronal precursors (NPs) would be useful for discovering new effective drugs to treat cerebral ischemic stroke.

Survival and directed differentiation of progenitor cells has been shown to play an important role in regeneration of damaged tissues in different organs including brain [40]. Neurological outcome in tPA knockout mice and intranasal administration of tPA demonstrated that it promotes neurological recovery after cerebral ischemia by stimulating axonal growth via epidermal growth factor receptor signaling [41]. Vascular endothelial growth factor (VEGF) enhances both angiogenesis and neurogenesis and increases permeability of the blood-brain barrier. It was found that heparan sulfate significantly enhanced VEGF mediated angiogenesis in rat brain endothelial cell line and increased the proliferation and differentiation of primary neural progenitor cells via VEGF pathway [42]. In a rat model of cerebral ischemia, human trophoblast progenitor cells were administered intravenously 24 hours post ischemia, an enhanced expression of angiogenic and neurogenic factors and neuron-like differentiated cells in rat brain along with a significant decrease in infarct volumes was observed at day 3 or day 11 [43]. A non-randomized open label phase I study of autologous CD34+ selected stem/progenitor cell therapy in cerebral ischemic stroke patients with NIHSS≥8 and within 7 days of stroke onset showed improvement in mRS and NIHSS scores during 6 month follow-up [44]. Patients with chronic stroke and functional deficits having NIHSS ≥ 6 were transfused intravenously with mesenchymal stem cell over the 12-months of follow-up BI scores increased (P< 0.001) by 10.8±15.5 points [45]. It appears that an approach of using stem cells or progenitor cells may have promise in improving the outcome of patients with acute cerebral ischemic stroke.

Sovateltide (an ET_B_ receptor agonist) injected intravenously showed higher neuronal progenitor cells (NPCs) differentiation along with better mitochondrial morphology and biogenesis in the brain stroked rats. Exposure of cultured NPCs to hypoxia also showed higher NPC differentiation and maturation with sovateltide [46, 47]. ET_B_ receptors are vital for the development of neural crest-derived epidermal melanocytes and enteric neurons [19, 27, 28, 48, 49]. ET_B_ receptors are known to regulate the differentiation, proliferation and migration of neurons, melanocytes and glia of both the enteric and central nervous systems during pre- and post-natal development [5, 20, 23, 50]. ET_B_ receptor knock-out in rodents led to mortality within 4 weeks of birth [51, 52] and had craniofacial malformation [53, 54] and had increased apoptosis with a significantly decreased number of neural progenitor cells in the CNS. It has been demonstrated that ET_B_ receptors in the brain are over expressed at the time of birth and their expression decreases with maturity of the brain [21, 55]. We have successfully used sovateltide as a novel pharmacological tool to activate a regenerative response in the ischemic brain by stimulating ET_B_ receptors.

Stimulation of ET_B_ receptors by sovateltide facilitated restoration of cerebral blood flow and improvement in neurological and motor function after stroke [24, 25, 29, 30]. Sovateltide treated animals showed strong evidence of neural tissue repair and regeneration including decrease in infarct volume and oxidative stress, increased pro-angiogenic, pro-survival and antiapoptotic markers as well as number of proliferating cells [24, 25, 29, 30, 46, 47]. A safe and maximum tolerated dose of sovateltide was determined in the clinical phase I trial (CTRI/2016/11/007509) in healthy human volunteers [5]. In the present study we are reporting results of an exploratory phase II (NCT04046484, CTRI/2017/11/010654) study in ischemic stroke patients.

Primary objective of the study was to evaluate the safety and tolerability of sovateltide. Key secondary objectives included efficacy measurements of neurological improvements by NIHSS, mRS and BI scales and quality of life assessments by Euro Qol and stroke-specific quality of life (SSQoL). Sovateltide was well tolerated and every patient received, without any resistance, all the 9 doses during treatment. Sovateltide treatment was initiated between 8 and 24 hours after the onset of stroke. Sovateltide treatment resulted in a significantly quicker recovery as measured by improvements in neurological outcomes in mRS and BI scales on day 6 compared to day 1. Moreover, sovateltide increased the frequency of favorable outcomes in all scales at 3 months. An improvement of ≥2 points in mRS was observed in 60% and 40% patients in sovateltide and saline groups, respectively. Similarly, BI improvement of ≥40 points was 64% and 36% in sovateltide and saline groups, respectively. Number of patients with complete recovery achieving NIHSS score of 0 and BI of 100 were significantly more (p< 0.05) in sovateltide group compared to saline group. Sovateltide treatment resulted in improved Quality of Life as measured by EuroQoL and SS-QoL (stroke specific quality-of-life) on day 90. No drug related adverse events were reported. Results indicate a clear superiority of sovateltide over SOC resulting in better clinical outcome of patients with acute cerebral ischemic stroke.

Patients enrolled in this study were with mild to moderate degree of acute stroke and patients with lacunar stroke was excluded. Main reason for excluding lacunar stroke was to exclude extreme variability in the assessment of neurological outcome in such strokes. Since this was a study using a potential first-in-class drug product we took this approach in order to have a proper comparison with limited number of patients. All the patients (100 percent) in control and sovateltide groups at the time of enrollment had mRS score > 2 in both control and sovateltide groups. Further, in control group 83.33 percent (15 of 18) patients and in sovateltide group 88.89 percent (16 of 18) patients had NIHSS score of ≥6 and BI score of < 60. Stroke in most of the patients was of moderate in nature because the NIHSS score was 9.17 ± 0.89 in control and 9.72 ± 0.94 in sovateltide groups. Similarly, mRS was 3.72 ± 0.11 in control and 3.78 ± 0.10 in sovateltide groups and BI was 35.56 ± 5.60 in control and 32.22 ± 4.75 in sovateltide groups. The most important clinical endpoint recognized with the major regulatory agencies is the mRS at 90 days and mRS was 1.71 ± 0.41 in control and 0.88 ± 0.26 in sovateltide groups at 90 days.

There is an indirect supporting evidence of reduced number of nonfatal strokes in the SONAR trial conducted with a selective ET_A_ receptor antagonist (leaving ET_B_ receptors unblocked) in type 2 diabetic patients with chronic kidney disease [56] having significantly greater risk of cardiovascular accidents [57]. Unblocked ET_B_ receptors can be stimulated by endogenous circulating ET-1 to produce neuroprotective effect, because ET_A_ selective antagonism produces an overstimulation of ET_B_ receptors [58] and that ET-3 (more selective for ET_B_ receptors) dilates cerebral blood vessels which was enhanced by ET_A_ receptor antagonist BQ123 [59]. These findings augment our argument that sovateltide mediates brain regeneration and repair after ischemic stroke in the adult brain. Further confirmation is provided by evidence that selective blockade of ET_B_ receptors exacerbates ischemic brain damage [19].

Limitation of this study is that it was conducted in small number of patients and was conducted in one country. This was an exploratory phase II study because action of sovateltide is a completely new approach to treat stroke. Promising results of this study have led to an efficacy study in a larger cohort of ischemic stroke patients and sovateltide is currently being tested in a multicentric, randomized, blinded, controlled efficacy clinical trial phase III (NCT04047563). Another limitation is that more female patients were enrolled in control group which could affect the outcome. It has been reported that age-adjusted mortality rates due to stroke are higher for men than women, however, women surviving stroke have less favorable outcomes than men [60]. Although, results of the present study suggest that sovateltide treatment can be initiated within a window of 24 hours after the onset of stroke, however, experimental studies in our laboratory indicate that if sovateltide treatment is initiated within 2–6 hours after the onset of stroke the neurological outcome is better. Hence, sovateltide treatment if initiated quickly after the onset of stroke a greater improvement in neurological outcome is possible. In the present study number of patients that received treatment after 20 hours of onset of stroke were 22.22% in the control group compared to 44.45% in sovateltide group. A greater number of patients received delayed treatment in sovateltide compared to control group.

This is the first clinical study where intravenous administration of sovateltide in patients with acute cerebral ischemic stroke showed an improvement in neurological outcome. Sovateltide induced stimulation of ET_B_ receptors has multimodal action aiming to prevent ischemic damage with restoration of blood flow, protection from apoptosis and differentiating adult neuronal progenitor cells to promote neural cell regeneration after stroke.

## Data Availability

The data that support the findings of this study are available upon reasonable request from the corresponding author. The data are not publicly available because of restrictions and containing information that could compromise the privacy of patients.

## Acknowledgments

We also acknowledge Pharmazz Inc. Willowbrook, IL, USA for funding this study.

## Authors contributions

Conceptualization: AG; Data curation: NA, DV, UKM, BP, DJ, JP and RB; Formal analysis: AG; Funding acquisition: AG; Investigation: NA, DV, UKM, BP, DJ, JP and RB; Methodology: AG, NA, DV, UKM, BP, DJ, JP and RB; Project administration: AG, NA, DV, UKM, BP, DJ, JP and RB; Visualization: NA, DV, UKM, BP, DJ, JP and RB; Writing–original draft: AG; Writing–review & editing: AG, NA, DV, UKM, BP, DJ, JP and RB

## Clinical Trial

The study protocol (PMZ-01 Version 2.0/April 18, 2016) was approved by the Drugs Controller General of India (DCGI), Directorate General of Health Services, Ministry of Health and Family Welfare, Government of India and Institutional Ethics Committee of each of 6 sites reviewed and approved the study protocol before the site was initiated. The study was registered at the Clinical Trials Registry, India (CTRI/2017/11/010654) and NCT04046484.

